# Evaluation of Complications Associated with Hypertonic Saline versus Mannitol in the Management of Intracranial Hypertension and Their Mitigation Strategies: A Systematic Review

**DOI:** 10.1101/2025.04.25.25326424

**Authors:** Mahsa Hemmati, Hesan Abbasi, Amirmohammad Bahri, Madi Fallah, Shahrzad Sharifzadeh, Sayeh Oveisi, Saeed Oraee Yazdani, Alireza Zali, Seyedpouzhia shojaei, Farzan Fahim

## Abstract

**Title:** Evaluation of Complications Associated with Hypertonic Saline versus Mannitol in the Management of Intracranial Hypertension and Their Mitigation Strategies: A Systematic Review

**Background:** Intracranial hypertension (ICH) worsens outcomes in patients with traumatic brain injury (TBI) or post-neurosurgical conditions. Hyperosmolar agents, particularly mannitol and hypertonic saline (HTS), are widely used to reduce intracranial pressure (ICP), but their safety profiles remain debated. This systematic review aimed to compare complications and mitigation strategies associated with mannitol and HTS in managing elevated ICP.

**Methods:** Eligible studies included randomized controlled trials (RCTs), cohort, and case-control studies from 2020 onward that reported complications related to mannitol or HTS in TBI or post-craniotomy patients. A comprehensive search was conducted in PubMed, Scopus, and Web of Science (last search February 17, 2025). Risk of bias was assessed using the Cochrane ROB2 and ROBINS-I tools. Due to heterogeneity in design and outcomes, a narrative synthesis was performed.

**Results:** Seven studies (3 RCTs, 4 observational) involving 14,367 patients were included. HTS was associated with fewer complications including reduced risks of electrolyte imbalance, acute kidney injury (AKI), and hemodynamic instability compared to mannitol. Mannitol was linked to hyponatremia, hypotension, and transient reductions in cardiac output, particularly in elderly or renally impaired patients. Mortality and ICU stay duration were generally comparable, though HTS showed potential ICU survival benefit. Evidence certainty ranged from low to moderate, with risk of bias and small sample sizes affecting confidence.

**Discussion:** HTS may offer a safer alternative to mannitol in patients at risk of renal or cardiac complications, with similar efficacy in ICP control. Mannitol remains effective but requires intensive monitoring. These findings support tailored use based on patient comorbidities and hemodynamic stability. For Iranian neurosurgical centers, where ICU bed shortages and cost constrains are prevalent, HTS’s renal safety profile and reduced cardiac strain in elderly patients could optimize resource utilization, a consideration absent in Western guidelines.

**Funding:** No external funding was declared for this review.

## Introduction

Intracranial hypertension, defined as an intracranial pressure (ICP) >20 mmHg for a period of more than 5 min, worsens neurologic outcome in traumatic brain injury (TBI).[1] Elevation of ICP to 20 mm Hg or greater can result in impaired brain perfusion, poor neurological outcome, and mortality.[2] In one review, the incidence of death was 18.4% for patients with an ICP <20 mm Hg but rose to 55.6% for those with an ICP greater than 40 mm Hg.[2] Brain has a very limited ability to compensate for hemorrhage, swelling, oedema, or mass effects due to the invariant constraints of the cranial vault.[3] Currently available medical treatments for raised intracranial pressure include hyperosmolar therapy, sedation and paralysis, hyperventilation, hypothermia, steroids and surgical intervention.[3] Mannitol (MTL) and hypertonic saline (HTS) are hyperosmolar solutions commonly used to reduce intracranial pressure (ICP) and brain volume reduction during brain surgery or critical care unit admissions.[4] Several clinical trials have compared the effects of HTS and mannitol on brain relaxation and ICP reduction in surgical and intensive care settings, and their results indicate that HTS is at least as effective as, if not superior to, mannitol in the treatment of increased ICP.[4] Mannitol represents the most commonly used osmotic agent, and numerous studies show its beneficial effect on both physiological parameters and neurological outcomes.[5] hypertonic saline has drawn increasing attention over the past two decades, with several studies suggesting a more potent effect of HTS in comparison with MTL.[5] Boluses of mannitol are often administered; however, its adverse effects include hypotension, electrolyte imbalance, and exacerbation of cerebral edema.[6] The initial increase in volume expansion can also result in pulmonary edema, which is concerning for patients with pre-existing cardiac or renal disease.[7] Adverse effects with HTS include pulmonary edema, acute renal failure (less likely compared with mannitol), and, although rare, possibly inducing central pontine myelinolysis by increasing serum sodium levels too quickly in the setting of baseline patient’s hyponatremia.[7] Despite their potential impact on mortality and neurologic outcome, limited studies have examined the risk factors for these complications in TBI patients.[8] Also comprehensive analysis of the complications associated with these interventions in the post-neurosurgical setting is still lacking. This systematic review aims to assess and compare the complications associated with mannitol and hypertonic saline in post-neurosurgical patients to determine their relative safety and efficacy.

## Methodology

This study was conducted as a systematic review based on the PRISMA (Preferred Reporting Items for Systematic Reviews and Meta-Analyses) guidelines.[9]

### Eligibility Criteria

Inclusion criteria: Studies meeting the following criteria were included in this systematic review: Randomized Controlled Trials (RCTs), Cohort Studies and case control studies from 2020 onwards examining the outcomes of hypertonic saline or mannitol use and their complications in patients having traumatic brain injury (TBI) or have undergone neurosurgical procedures for tumor resection. Studies were grouped by design (RCTs vs. observational) and population (TBI vs. craniotomy).

Our inclusion criteria prioritized post-2020 studies to reflect contemporary ICP management protocols, particularly those adapted during the COVID-19 pandemic. This focus inherently limited the number of eligible studies but ensured clinical relevance to current practice

Exclusion Criteria: case series, non-human studies, conference abstracts, book chapters and studies published in languages other than English were excluded.

### Search Strategy

A systematic search was conducted in the databases PubMed, Scopus, and Web of Science on February 17,2025. Additionally, reference lists of relevant articles were reviewed to identify more studies.

Keywords used in the search included “Traumatic Brain Injury (TBI)”, “Neurosurgery”, “mannitol”, “hypertonic saline”, “intracranial pressure (ICP)”, “complications”.

### Study Selection Process

The titles, abstracts, and full texts of the identified studies were carefully reviewed by two reviewers independently using Rayyan and based on predefined eligibility criteria. Any uncertainties during the process were discussed and resolved among the reviewers.

### Data collection

Two reviewers independently extracted data from each included study using a standardized data extraction form.

### Data items

We sought data on all reported complications related to the administration of hypertonic saline or mannitol in the management of intracranial hypertension. Primary outcomes included the incidence, type, and severity of adverse events such as electrolyte imbalance, kidney dysfunction, hemodynamic instability, cardiac function and osmotic demyelination syndrome. Where available, we also collected data on mortality, neurologic outcomes, and duration of ICU/hospital stay as secondary outcomes. All results that were compatible with each outcome domain across different studies, time points, and measurement methods were included.

In addition to outcomes, we extracted information on study characteristics (first author, year of publication, country, study design, sample size, inclusion and exclusion criteria), participant characteristics (age, sex, diagnosis, baseline ICP), and intervention details (type, concentration, dosage, and route of administration of mannitol or hypertonic saline). In cases of missing or unclear information, we referred to supplementary material or attempted to contact study authors. If this was not possible, we noted the missing data and made no assumptions unless otherwise stated in the original study.

### Risk of Bias Assessment

Potential bias in the included studies was assessed by two reviewers independently using Cochrane Risk of Bias Tool (ROB2) for RCTs and Cochrane Risk of Bias in Non-randomized Studies - of Interventions (ROBINS-I) for observational studies.

### Effect Measures

For each outcome domain, we recorded the effect measures reported by the included studies. Binary outcomes (e.g., presence of acute kidney injury, mortality) were summarized using odds ratios (ORs) with 95% confidence intervals (CIs) where reported. Continuous outcomes (e.g., serum sodium levels, urine output, hemodynamic parameters, ICU/hospital stay duration) were extracted as mean differences (MD) or median values with interquartile ranges (IQRs) when available. For outcomes such as hypotension or electrolyte imbalance, p-values were used to reflect statistical significance when effect size measures were not reported. Due to heterogeneity in study designs and outcome reporting, a meta-analysis was not performed, and all effect measures were presented descriptively as part of a narrative synthesis.

### Data synthesis

Eligibility for synthesis was determined by reviewing study characteristics, including interventions used (type, dose, and concentration of hyperosmolar therapy) and outcome domains reported. Studies were considered eligible for synthesis if they directly compared hypertonic saline and mannitol and reported at least one relevant complication or adverse effect.

No imputation or conversion of effect estimates was performed. When summary statistics such as standard deviations or confidence intervals were missing, results were reported descriptively using available p-values or narrative summaries. Incomplete data were acknowledged, and no statistical conversions were applied due to the small number of studies and heterogeneity in reporting formats.

Results from individual studies were tabulated in a summary of findings table (Table 3), which included the type of study, number of patients, intervention and comparator, reported outcomes, effect, and the GRADE certainty rating. Additionally, results were presented narratively within outcome domains in the Results section. Risk of bias assessments were visually summarized in Figures 1 and 2 using the ROB2 and ROBINS- I tools, respectively.

**Fig. 1.**
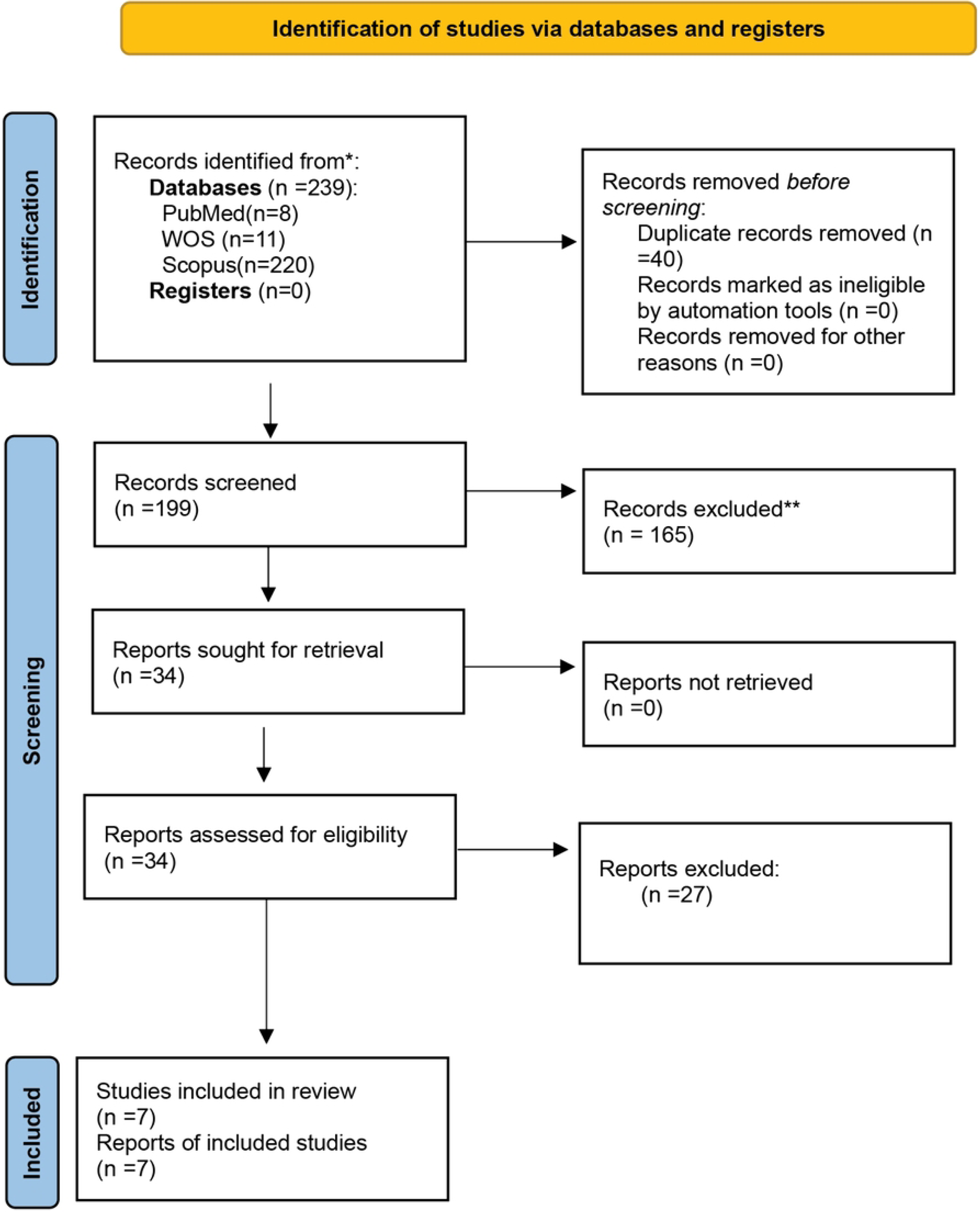
Preferred Reporting Items for Systematic Reviews (PRISMA) flow chart. showing studies identified, screened, and included in the systematic review

**Fig. 2.**
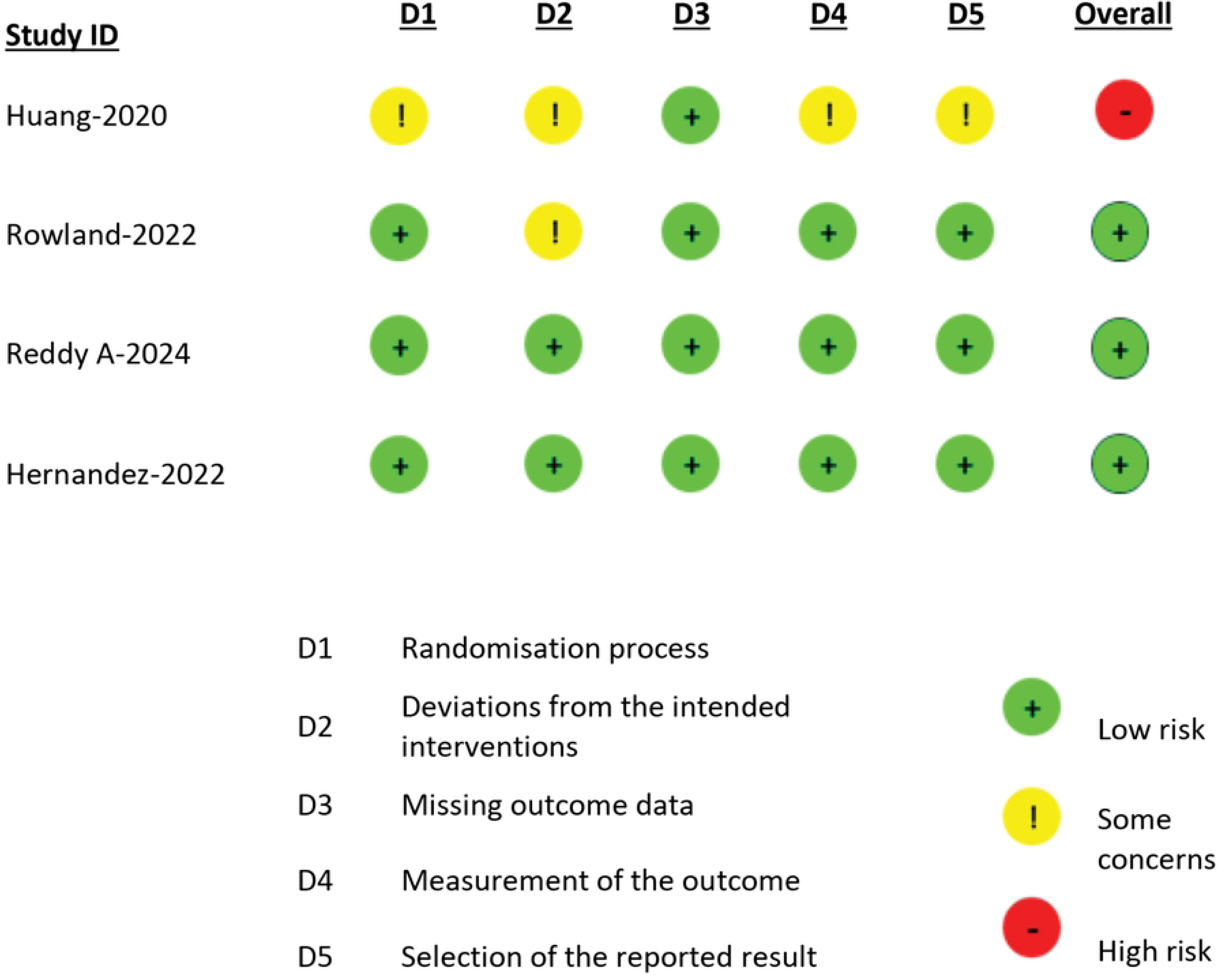
Risk of bias summary for RCT studies. (ROB2)

Given substantial heterogeneity across studies in terms of design, interventions, dosing regimens, populations, and outcome measures, a quantitative meta-analysis was not feasible. Therefore, we employed a narrative synthesis approach. This allowed for qualitative comparison across studies, highlighting consistency, directionality, and strength of associations where available. This method was chosen to preserve context and clinical detail that might otherwise be lost in statistical pooling.

Due to the limited number of studies and lack of standardized reporting, formal heterogeneity exploration such as subgroup analysis or meta-regression was not conducted. However, we qualitatively considered differences in patient populations, intervention protocols, and outcome definitions as potential sources of variation in the findings.

No formal sensitivity analyses were performed due to the narrative nature of the synthesis and the small number of studies included. However, we noted variations in study quality, sample size, and blinding in the risk of bias assessment and considered these factors when interpreting the strength and reliability of the evidence.

### Reporting Bias Assessment

Given the limited number of studies included and the use of a narrative synthesis rather than a meta-analysis, no formal statistical methods (e.g., funnel plots or regression-based tests) were applied to assess reporting bias. Any inconsistencies or missing outcome data were considered in the overall risk of bias and GRADE certainty assessments.

### Certainty of Evidence

The Grading of Recommendations Assessment, Development, and Evaluation (GRADE) approach was used to assess the overall certainty of the evidence[10].

## Results

A total of 239 studies were initially identified. After removing 40 duplicate studies, 165 articles were excluded following title and abstract screening, and 27 additional articles were excluded after full-text review. Ultimately, 7 studies were included in the final analysis: 3 randomized controlled trials (RCTs) and 4 observational studies. (Fig 1)

No studies were identified that appeared to meet inclusion criteria but were ultimately excluded based on subjective judgment or unclear eligibility. All full-text exclusions were based on clearly predefined criteria, and no further justification was required for any specific study.

### Study characteristics

The characteristics of the included RCTs are summarized in Table 1, while those of observational studies are in Table 2. 3. RCTs were included with a total of 174 patients, 2 of which utilized double- blinding. All RCTs patients were adult (>16 years), but one study focused on elderly patients (>64 years). Two RCTs included patients undergoing supratentorial craniotomy, while the other one focused on traumatic brain injury (TBI) patients.

**Table 1.** Characteristics of included RCT studies.

| Author | Method | Participants | intervention | comparator | Primary Outcome | Secondary Outcome |
| --- | --- | --- | --- | --- | --- | --- |
| **Hernandez et al. [11]** | Single center, double blinded RCT | Adult patients scheduled to undergo supratentorial craniotomy for brain tumor<br>n=60 | 5 mL/kg of 20% mannitol (1 g/kg, osmolality = 1.098 mOsm/L) administered through a central line for 15 min using an infusion pump | 5 mL/kg of 3% HTS (osmolality = 1.024 mOsm/L) administered through a central line for 15 min using an infusion pump | brain relaxation, assessed using a four-point scale by the surgeon | postoperative complications, perioperative outcomes, ICU and hospital length of stay, and mortality at 30 days |
| **Reddy A et al. [12]** | Single center, double blinded RCT | Elderly patients (age >65 years) undergoing supratentorial craniotomy With raised ICP<br>n=31 | 5 ml/kg of 20% mannitol infused over 20 minutes before dural opening using a 50-cc syringe and an infusion pump | 5.35 ml/kg of 3% HTS solution infused over 20 minutes before dural opening using a 50-cc syringe and an infusion pump | LVOT-VTI Used as an indicator of cardiac function and systemic hemodynamics. | Cardiac Output (CO), Blood Pressure, Total Fluid Intake and Urine Output, Brain Relaxation Score, Serum Electrolyte Levels |
| **Huang et al. [13]** | Single center RCT | eligible patients at least 18 years old, with severe TBI (GCS < 8) at admission, and with an ICP intraparenchymal monitor<br>n=83 | 20% mannitol (administered at a dosage of 2ml/kg) infused as a bolus via a central venous catheter over 15 minutes | 10% HTS (administered at a dosage of 0.63 ml/kg) infused as a bolus via a central venous catheter over 15 minutes | ICP reduction, Time to lowest ICP, Duration of effect | CPP changes, MAP and CVP variations Serum sodium and plasma osmolality changes |

ICP, intracranial pressure; HTS, hypertonic saline; LVOT-VTI, left ventricular outflow tract velocity time integral; TBI, traumatic brain injury; GCS, Glasgow coma scale; CPP, cerebral perfusion pressure; MAP, mean arterial pressure; CVP, central venous pressure.

**Table 2.** Characteristics of included observational studies.

| Author | Method | Participants | intervention | comparator | Primary Outcome | Secondary Outcome |
| --- | --- | --- | --- | --- | --- | --- |
| **Kochanek et al. [14]** | observational cohort comparative effectiveness research analysis | Age<18 years, diagnosis of TBI, ICP monitor used as part of the standard care, and GCS score ≤8 at time of monitor placement. N=787 | administration of mannitol in 1 hour of data collection | administration of HTS in 1 hour of data collection | ICP reduction | CPP changes |
| **Codorniu et al. [15]** | A retrospective observational cohort study | patients aged≥16 years with suspected TBI (GCS≤12) associated with a mydriasis n=1417 | Mannitol (dosage and administration were not provided) | HTS (dosage and administration were not provided) | ICU all-cause mortality | Early death, pupillary abnormality after, osmotherapy, Length of mechanical ventilation, Length of ICU stay, Length of hospital stay |
| **Li J et al. [16]** | single-center, retrospective, matched case-control study | Patients undergoing brain tumor resections. Age≥18 n=9933 | preoperative administration of mannitol | the control group of patients who did not develop AKI | association between mannitol infusion and postoperative AKI | associations between other perioperative factors and postoperative AKI (Tumor characteristics, use of NSAIDs, BP) |
| **Veen et al. [17]** | prospective multicenter cohort study | Patients over 16 years old with diagnosed TBI within the past 24 hours and with indication for a head CT n=2056 | Mannitol | HTS | influence of center on HOA preference: mannitol, HTS, or both | the association between the preference for a HOA on outcomes, defined as ICU mortality and the GOSE, assessed at 6 months after in |

TBI, traumatic brain injury; ICP, intracranial pressure; GCS, Glasgow Coma Scale; CT, computed tomography; AKI, acute kidney injury; CPP, cerebral perfusion pressure; BP, blood pressure; HOA, hyperosmolar agent; NSAID, non-steroidal anti- inflammatory drug; GOSE, Glasgow outcome scale-extended.

In all RCTs, the intervention groups received mannitol, and the comparator groups received hypertonic saline (HTS) at specific doses and timings, ensuring similar conditions for administration. (Table1) However, concentration and administration methods varied across studies.

Among the 4 observational studies, 3 were cohort studies, and one was a case-control study. One observational study focused on pediatric patients (<18 years), while the others included adult patients. One study examined patients undergoing brain tumor resection, while the others included TBI patients. The total number of patients in observational studies was 14,193.

### Risk if bias assessment

Details on risk of bias assessment for RCTs are available in Fig 2 and for observational studies are in Fig 3. RCT studies were assessed using Cochrane risk of bias 2 tool (ROB2). 2 studies were double blinded and had low risks of bias in all domains. The other study had a high risk of bias due to lack of blinding, absence of allocation concealment details, and missing a prespecified analysis plan. The study risk of bias was concerning in randomization, deviation from the intended intervention, outcome measurement, and selection of reported results, making it high risk overall.

**Fig. 3.**
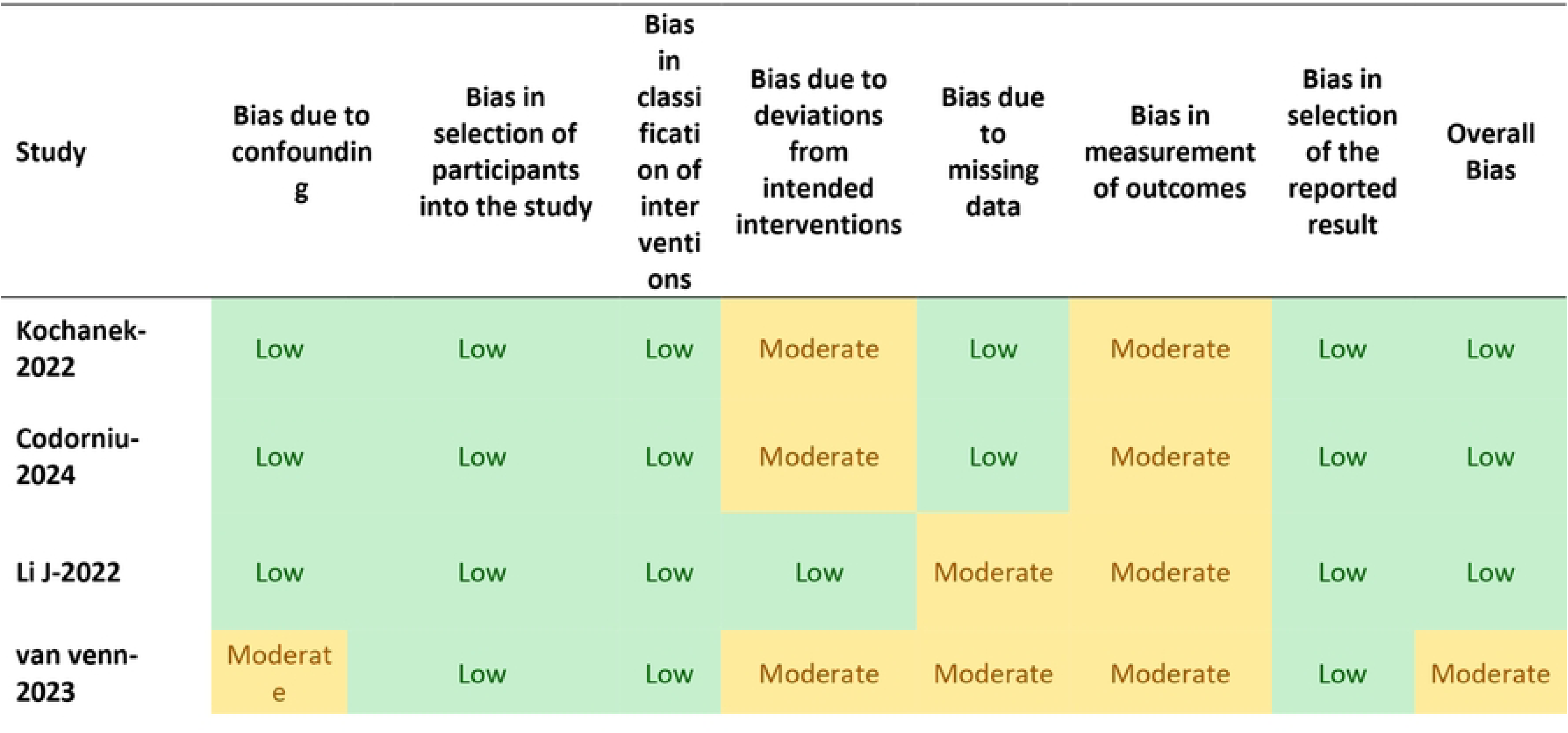
Risk of bias summary for observational studies. (ROBIN-I)

For observational studies Risk of Bias in Non-randomized Studies - of Interventions (ROBIN-I) tool was used. Most studies had a moderate risk of bias in the deviation from intended intervention domain, due to variations in dosage and administration methods or lack of reporting on these details. 3 studies had moderate concerns in the outcome measurement domain, as assessors were aware of the interventions Two studies had a high risk of bias due to missing data.The overall risk of bias was low for all studies except one, which had a high risk due to incomplete reporting.

### Results of individual studies

For each study, a summary of statistics for all reported outcomes is presented in Table 3.

**Table 3.** Summary results of individual studies.

| study | Outcomes | measure | Mannitol Group (n, Mean/Incidence ± SD) | HTS Group (n, Mean/Incidence ± SD) | p-value |
| --- | --- | --- | --- | --- | --- |
| **Hernandez et al.[11]** | Electrolyte imbalance | Urine output | 1612±602 (n=30) | 1236±738 (n=30) | 0.037 |
| | Hemodynamic stability | MAP | 71 ± 8 (n=30) | 73 ± 10 (n=30) | 0.586 |
| | | CVP | 7 ± 3 (n=30) | 7 ± 2 (n=30) | 0.574 |
|  | Hospital length of stay | PACU stay | 20.00 (n=30) | 19.00 (n=30) | 0.647 |
|  |  | Hospital stay | 8.00 (n=30) | 8.00 (n=30) | 0.966 |
|  | Mortality and morbidity | Average complications incidence | 9 (n=30) | 9.125 (n=30) | 0.95 |
| **Reddy et al.[12]** | Electrolyte imbalance | Urine output | 401.07±41.52 (n=14) | 224.29±24.09 (n=14) | <0.001 |
|  |  | Serum sodium | 137.87 (n=14) | 143.07 (n=14) | <0.001 |
|  | Hemodynamic instability | MAP, DBP |  |  | > 0.05 |
|  |  | Hypotensive episodes | Higher incidence | Lower incidence | 0.009 |
| | Cardiac function | LVOT-VTI (cm) | 16.76±1.81 (n=14) | 20.78±1.87 (n=14) | 0.001 |
| | | Cardiac Output (ml/minute) | 3863.16±845.87 (n=14) | 4745.59±1209.33 (n=14) | 0.034 |
| **Huang et al.[13]** | Hemodynamic stability | MAP | 94.2 ± 7.1 (n=236) | 93.9 ± 8.3 (n=221) | > 0.05 |
| | | CVP | 8.4 ± 2.5 (n=236) | 8.6 ± 2.9 (n=221) | > 0.05 |
|  | Osmotic demyelination syndrome | No cases of CPM |  |  |  |
|  | Kidney dysfunction | 1 reported case of severe kidney dysfunction |  |  |  |
| **Kochanek et al.[14]** | Electrolyte imbalance | maximum serum osmolarity observed | 331.66 ± 23.82 (n=74) | 312.90 ± 39.76 (n=74) | <0.001 |
| **Codorniu et al. [15]** | Hemodynamic stability (after PSM) | Worst pulsatility index on transcranial | 1.4 (1.1–2) (n=720) | 1.3 (1.1–1.8) (n=240) | 0.22 |
|  |  | Worst diastolic velocity on transcranial | 20 (13–27) (n=720) | 21 (18–30) (n=240) | 0.52 |
|  | length of stay (after PSM) | ICU length of stay | 5 (2–18) (n=720) | 7 (2–25) (n=240) | 0.72 |
|  |  | Hospital length of stay | 7 (2–32) (n=720) | 10 (2–37) (n=240) | 0.35 |
|  | Mortality (after PSM) | ICU all-cause mortality | 389 (54) (n=720) | 107 (45) (n=240) | 0.014 |
|  |  | Early death | 166 (23) (n=720) | 57 (24) (n=240) | 0.86 |
| **Li et al. [16]** | Kidney dysfunction | Risk factors of postoperative AKI |  |  | 0.034 |
| **Van Veen et al. [17]** | Mortality and morbidity | ICU mortality (%) | 28 (19.3) (n=142) | 58 (20.2) (n=287) | 0.011 |

MAP, mean arterial pressure; CVP, central venous pressure; PACU, post-anesthesia care unit; DBP, diastolic blood pressure; LVOT-VTI, left ventricular outflow tract velocity time integral; CPM, Central Pontine Myelinolysis; PSM, propensity score matching; ICU, intensive care unit; AKI, acute kidney injury;

### Narrative Synthesis

#### Electrolyte Imbalance

Mannitol was consistently associated with a stronger diuretic effect (P=0.037, n=60), leading to decreased serum sodium due to water loss and dilution. One RCT found that this effect resulted in a significant drop in serum sodium, which gradually normalized over time.[11] Another RCT reported that urine output was significantly higher in the mannitol group (P < 0.001, n=31), whereas the HTS group exhibited transient increases in serum sodium and chloride, which normalized within 24 hours.[12]

Observational studies suggested that despite HTS delivering nearly twice the osmolar load compared to mannitol, peak serum osmolarity was not higher in the HTS group(n=787)[14]. This could indicate a lower risk of hypernatremia or osmotic complications with HTS, though clinical consequences such as renal failure or seizures were not explicitly addressed.

These findings are consistent across two well-conducted RCTs and two large observational studies. The RCTs had low risk of bias, while observational studies had moderate risk due to variation in outcome measurement and confounding. Together, these studies suggest that both therapies can cause electrolyte imbalances, necessitating close monitoring of serum sodium and osmolarity during hyperosmolar therapy.

#### Hemodynamic Parameters

Mannitol’s potent diuretic effect raises concerns about its impact on hemodynamic stability. One RCT noted that HTS did not significantly reduce the total intravenous fluid volume required to maintain stable mean arterial pressure (MAP) and central venous pressure (CVP). [11]

Another RCT reports that MAP and CVP varied slightly after osmotherapy, but there was no significant difference between the 2 groups (P>.05).[13]

However, another RCT reported a higher incidence of hypotensive episodes in the mannitol group, suggesting a potential risk of hypovolemia (P=0.009). This study state that there was no significant difference in MAP and DBP between the 2 groups but further analysis within group M revealed that SBP, DBP, and MAP showed a significant increase at 15 mins ( P= 0.014, <0.001, and < 0.001 respectively) followed by a significant fall at 45, 60, and 90 minutes as compared to the baseline and concludes that the systemic hemodynamics were better maintained in group HS.[12]

An observational study found no significant differences in worst pulsatility index or diastolic velocity on transcranial Doppler(P=0.22, P=0.52).[15]

This synthesis draws from three RCTs and one observational study, all of which enrolled adult patients undergoing craniotomy or TBI management. While RCTs were generally low in bias, one lacked full blinding and protocol transparency. The observational study used propensity score matching(PSM) to improve comparability, reducing but not eliminating confounding.

These findings imply that while mannitol may pose a higher risk of hypovolemia and hemodynamic instability, the statistical differences in hemodynamic outcomes remain inconclusive. Careful fluid management strategies are essential, especially in patients vulnerable to hypotension.

#### Osmotic Demyelination syndrome

Osmotic demyelination syndrome (ODS), including central pontine myelinolysis (CPM), was not explicitly reported in any included studies. One RCT suggested that alternating between HTS and mannitol in the same patient could mitigate the risk of ODS.[13]

Another RCT indicated that complications such as osmotic demyelination were linked to prolonged infusion or high doses of HTS, emphasizing the importance of controlled infusion over at least 20 minutes to reduce this risk.[12]

#### Kidney Dysfunction

Mannitol has been historically associated with acute kidney injury (AKI), particularly at higher doses. One RCT documented one case of severe kidney dysfunction following HTS administration, but observational data suggested that HTS was associated with a lower risk of AKI during ICU stays.[13]

One observational study identified preoperative mannitol administration as an independent risk factor for postoperative AKI (OR 1.64; 95% CI, 1.04–2.60; P = 0.034), suggesting that patients receiving mannitol should have closely serum creatinine monitoring .intraoperative and postoperatively hypotension should be avoided to mitigate renal risk. This large cohort study mainly discuss preoperative risk factors associated with AKI and despite some concerns about outcomes measurement and missing data, it has low risk of bias [16]

#### Cardiac Function

One RCT found that in elderly patients undergoing supratentorial craniotomy, the mannitol group had a significant decrease in left ventricular outflow tract velocity-time integral (LVOT-VTI) at 45 and 60 minutes, compared to the HTS group.[12] This suggests that mannitol may negatively impact cardiac function in older patients, possibly due to its volume-depleting effects. No other studies explicitly examined cardiac function, highlighting a potential knowledge gap in this area. This outcome was reported in a single, double-blinded RCT with low risk of bias. However, as it is based on a highly specific population (elderly patients), generalizability is limited.

#### Hospital and ICU Length of Stay

The available evidence suggests that mannitol and HTS have no significant difference in hospital or ICU length of stay. One RCT reported similar PACU and hospital stays between groups after supratentorial brain tumor surgery(P=0.647).[11] An observational study similarly found no significant difference in ICU or hospital length of stay between mannitol and HTS groups.[15] Risk of bias was low in both the RCT and the observational study. The findings appear consistent.

#### Mortality and Morbidity

Mortality outcomes varied across studies. One RCT reported no significant differences in overall postoperative morbidity(P=0.966) with low risk of bias.[11] An observational study found lower ICU all-cause mortality in the HTS group (OR 0.68 (0.5–0.9), P = 0.014), but no difference in early mortality.(P=0.86).In this study PSM was used to reduce confounding and was assessed as low risk of bias [15] Another observational study also reports no significant difference in mortality between the two groups(OR = 1.0, CI = 0.4 – 2.2) but this study had moderate risk of bias du to some factors such as confounding and missing data.[17] While some evidence suggests a survival benefit with HTS in ICU settings, the overall mortality rate is mostly similar

### Certainty of evidence

The certainty of evidence ranged from low to moderate across most outcomes (Table 4). Common reasons for downgrading included risk of bias, inconsistency in effect estimates, and imprecision due to small sample sizes. While RCTs provided stronger evidence, variations in study design, intervention protocols, and outcome assessments contributed to uncertainty. Observational studies supplemented findings but were subject to potential confounding and publication bias. Further well-powered trials are needed to improve confidence in these results.

**Table 4.** Summary of Findings Table: Complications associated with Mannitol vs. Hypertonic Saline for ICP Management.

| Outcomes | No. of Studies | Study Type | Mannitol Group Findings | HTS Group Findings | Effect Estimate | Certainty of Evidence (GRADE) |
| --- | --- | --- | --- | --- | --- | --- |
| Electrolyte Imbalance | 3 RCTs, 1 Obs | RCTs, Cohort | Greater diuretic effect, transient hyponatremia, increased urine output | Transient hypernatremia & hyperchloremia, normalizing within 24h | P=0.037 (n=60), P<0.001 (n=31) | **High** ↑↑↑↑ (Consistent effect, multiple studies) |
| Hemodynamic Parameters | 3 RCTs, 1 Obs | RCTs, Cohort | Higher risk of hypotension in one RCT; others showed no difference in MAP/CVP | No significant impact on MAP/CVP | P=0.009 (hypotension in mannitol group) | **Moderate** ↑↑↑ (Possible dose-response with diuresis) |
| Osmotic Demyelination | 2 RCTs | RCTs | No cases reported, one RCT suggested alternating therapies may lower risk | No cases reported | No cases reported | **Low** ↑↑ (Risk may be underestimated due to underreporting) |
| Kidney Dysfunction | 1 RCT, 2 Obs | RCT, Cohort | Increased AKI risk (Mannitol linked to pre-op AKI, OR 1.64, P=0.034) | 1 RCT reported 1 severe AKI case with HTS | OR 1.64 (95% CI, 1.04–2.60), P=0.034 | **Moderate** ↑↑↑ (Large effect size in observational data) |
| Cardiac Function | 1 RCT | RCT | Decreased LVOT-VTI in elderly (suggesting reduced cardiac function) | No reported cardiac effects | Significant LVOT-VTI decrease (P<0.05) | **Moderate** ↑↑↑ (Large effect in elderly) |
| Hospital and ICU Stay | 1 RCT, 1 Obs | RCT, Cohort | No significant difference in PACU or hospital stay | No significant difference | P=0.647 (hospital stay) | **Low** ↑↑ (No strong evidence) |
| Mortality and Morbidity | RCT, 2 Obs | RCT, Cohort | No significant difference in morbidity | Lower ICU mortality in one study, but not others | OR 0.68 (0.5–0.9), P=0.014 (ICU mortality) | **Moderate** ↑↑↑ (HTS may benefit ICU mortality, but data is mixed) |

RCT randomized clinical trial, Obs observational studies, MAP mean arterial pressure, CVP central venous pressure, AKI acute kidney injury, LVOT-VTI left ventricular outflow tract velocity time integral, PACU post-anesthesia care unit

## Discussion

This study provides a comparative analysis of HTS and mannitol for managing elevated intracranial pressure (ICP), highlighting significant differences in their associated complications and management strategies. Mannitol demonstrated stronger diuretic effects, resulting in a higher incidence of electrolyte imbalances, particularly hyponatremia, necessitating close monitoring of serum sodium levels. Conversely, HTS showed a lower risk of hypernatremia and osmotic complications, even when delivering a higher osmolar load, which may reflect its relative safety in electrolyte management. Hemodynamic instability, including a higher incidence of hypotension, was more commonly observed with mannitol. In contrast, HTS generally maintained hemodynamic stability, making it a potentially safer option for patients at risk of hypotension.

Although complications such as osmotic demyelination were rare, they underscore the importance of using controlled infusion protocols and alternating therapies to minimize risks. Acute kidney dysfunction was observed more frequently with mannitol, particularly in perioperative settings, whereas HTS posed a comparatively lower risk of acute kidney injury. Cardiac function appeared to be negatively impacted by mannitol in elderly patients, an area warranting further RCTs. Notably, both agents had comparable effects on ICU and hospital lengths of stay, as well as overall morbidity rates. While HS demonstrated a potential survival benefit in ICU settings, mortality outcomes were largely similar across both groups.

These findings are consistent with existing literature that recognizes HTS as at least as effective as mannitol for reducing ICP, with a potentially better safety profile. However, the evidence certainty ranged from low to moderate, reflecting variations in study design, sample sizes, and outcome measures. This highlights the need for future research to address these limitations and improve evidence quality.

For Iranian neurosurgical centers, where ICU bed shortages and cost constrains are prevalent, HTS’s renal safety profile and reduced cardiac strain in elderly patients could optimize resource utilization, a consideration absent in western guidelines.

### Clinical pearls

1. Cardiac function in elderlies: mannitol is associated with reduced left ventricular outflow tract velocity time integral (LVOT-VTI) at 45-60 minutes post administration (p <0.05), suggesting transient impairment of cardiac function. HTS may be preferable in this population.
2. Electrolyte imbalance: while HTS transiently elevates serum sodium and chloride, which normalized within 24 hours. Conversely, mannitol requires frequent sodium monitoring due to hyponatremia risk (p< 0.037).
3. Renal dysfunction risks: preoperative mannitol administration increases postoperative AKI risk (OR 1.64;CI 95%; P<0.034), particularly at cumulative doses >1.5g/kg. serum creatinine should monitor for some days postoperatively.
4. Osmotic demyelination mitigation: controlled HTS infusion over 20 minutes may reduce osmotic demyelinating risk. No cases were reported in included studies, but theorical risks remain.
5. Mortality nuance: while HTS may reduce ICU all-cause mortality (OR:0.68; CI 95%, 0.5-0.9; P<0.014), no survival benefits were observed in early mortality or overall morbidity.
6. Risk of bias transparency: Figure 2-3(Risk of Bias assessment) demonstrates moderate to high methodological quality in RCTs but significant confounding risks in observational studies. These findings should be interpreted with caution in hemodynamically unstable patients.
7. GRADE certainly context: the moderate certainly for hemodynamics outcomes and low certainly for hospital stay (table 3) highlight the need for individualized therapy. Strongest evidence supports HTS for electrolyte stability (high certainly)
8. Practical protocol recommendation:

1. Initiate HTS at 3% (2-5 ml/kg over 20-30 mins).
2. Reserve mannitol (0.5-1 g/kg) for normotensive and normal renal function patients.
3. Monitor serum sodium closely post hypertonic solutions administration.

### Study Limitations and Implications

The study’s findings are limited by heterogeneity in the included studies, such as differences in patient populations, dosing protocols, and outcome definitions. These factors limit the generalizability of the results and underscore the need for standardized treatment protocols. The exclusion of non-English studies, while aimed at minimizing language bias, may have omitted relevant non-English speaking regions. This is particularly relevant for interventions like hypertonic saline, which may have unique utilization patterns in resource-limited setting. Furthermore, the small sample sizes in some RCTs and potential biases in observational studies reduce the reliability of the conclusions. Despite these limitations, the study has important implications for clinical practice. Given the comparable efficacy and differences in safety profiles, HS may be a preferred option in patients at higher risk of hemodynamic instability or acute kidney injury.

Mannitol, while effective, requires close monitoring of electrolyte and cardiac parameters, particularly in vulnerable populations such as the elderly.

### Future Directions

Further well-powered RCTs are needed to refine our understanding of the optimal dosing, timing, and patient selection for both HS and mannitol. Subgroup analyses based on factors such as age, comorbidities, and surgical context could provide more personalized insights. Additionally, research exploring long-term outcomes, including functional recovery and quality of life, would be valuable for guiding treatment decisions.

In summary, while both HS and mannitol remain essential tools in ICP management, their unique risk profiles necessitate individualized therapeutic strategies to optimize safety and efficac

## Conclusion

This study compares hypertonic saline (HTS) and Mannitol, focusing on complications and their management. Due to the stronger diuretic effects of Mannitol, it has a greater risk of electrolyte imbalance and hyponatremia. The use of Mannitol can also increase the risk of acute kidney injury (AKI) and has also been linked to a higher rate of hypotensive events, which indicates hemodynamic instability, Especially in elderlies. Rapid administration of high doses of HTS can increase the risk of Osmotic Demyelination Syndrome (ODS), which can be avoided with prolonged administration of lower doses. HTS appears to provide a more stable osmolar response with a potentially lower risk of AKI. Mortality and ICU stay outcomes were generally similar, though some evidence exists for survival benefit with HTS. With low to moderate certainty of evidence, further well-powered studies will be needed to provide definitive guidance. Individualized patient monitoring is still needed to optimize treatment effects.

For Iranian neurosurgical centers, where ICU bed shortages and cost constraints are prevalent, HTS’s renal safety profile and reduced cardiac strain in elderly patients could optimize resource utilization, a consideration absent in Western guidelines.

## Data Availability

All relevant data are within the manuscript and its Supporting Information files.

